# Key predictors of maternal mild depression and anxiety in low resource settings: A machine learning approach

**DOI:** 10.1101/2025.05.08.25327241

**Authors:** Faith Neema Benson, Rachel Odhiambo, Anthony K. Ngugi, Willie Brink, Akbar K. Waljee, Cheryl A. Moyer, Ji Zhu, Amina Abubakar

## Abstract

**Background:** Maternal mental health (MMH) disorders, particularly depression and anxiety, are major public health concerns in low- and middle-income countries (LMICs). In sub-Saharan Africa (SSA), their prevalence remains alarmingly high, yet identifying key predictors is challenging due to the limitations of traditional statistical methods in capturing complex risk factor interactions.

**Objective:** This study aimed to identify key predictors of maternal depression and anxiety using machine learning (ML) techniques in a low-resource setting. Additionally, the study sought to determine the prevalence of both conditions within the population.

**Methods:** A cross-sectional study was conducted in Kaloleni and Rabai sub-counties in Kilifi, Kenya. Data were collected from 1,995 mothers of singleton children aged 0 to 6 months between March 2023 and March 2024. Depression and anxiety symptoms were assessed using the Patient Health Questionnaire-9 (PHQ-9) and Generalized Anxiety Disorder-7 (GAD-7) scales, respectively. Additional data on sociodemographic factors, food insecurity, health history, nutrition, and socioeconomic status were collected as potential predictive features. Three supervised ML models including Random Forest (RF), Logistic Regression (LR) with L2 regularisation (Ridge), and Extreme Gradient Boosting (XGBoost) were applied to predict depression and anxiety symptoms. Model performance was evaluated using metrics including the area under the receiver operating characteristic curve (AUC), accuracy, sensitivity, and specificity. Feature selection and model interpretability were performed using SHapley Additive exPlanations (SHAP) values.

**Results:** The prevalence of maternal depression and anxiety symptoms was 15.14% and 8.67%, respectively. Although there were no statistically significant differences in prediction performance among the three models, the RF model showed a slightly better performance in predicting anxiety symptoms, with an AUC of 78.9%, accuracy of 72.9%, sensitivity of 74.3%, and specificity of 72.8%. LR performed slightly better in predicting depression symptoms, achieving an AUC of 72.4%, accuracy of 69.9%, sensitivity of 63.3%, and specificity of 71.1%. Food insecurity emerged as a key predictor for both outcomes, followed by low wealth index, increased maternal age, lower body mass index (BMI), higher number of children, and pregnancy complications.

**Conclusion:** This study highlights the key predictors of maternal depression and anxiety in a low-resource setting, with food insecurity emerging as a critical predictor. Other key predictors included low wealth index, increased maternal age, lower BMI, higher number of children, and pregnancy complications. ML models showed potential for identifying high-risk individuals and supporting targeted interventions. However, limitations such as imbalanced data, reporting bias, and the cross-sectional design may affect generalizability. Larger, longitudinal studies are needed to validate these findings and enhance predictive performance.

## Introduction

Maternal mental health (MMH) challenges during pregnancy and postpartum are a significant global public health issue, affecting approximately 10% of pregnant women and 13% of postpartum mothers (1-3). Depression and anxiety are the most common MMH disorders (4). The burden of maternal depression and anxiety is particularly high in low- and middle-income countries (LMICs), where prevalence estimates are two to three times higher than in high-income countries (5-7). In LMICs, maternal depression affects 10% to 25% of women, while anxiety ranges from 8% to 16% during pregnancy and postpartum (6, 8). In Sub-Saharan Africa (SSA), these rates are even more alarming, with depression prevalence ranging from 10% to 70% and anxiety from 10% to 50% (9-13).

Mothers experiencing depression and anxiety disorders often struggle to function effectively in daily life and, in severe cases, may have suicidal thoughts (3). These conditions not only compromise maternal well-being but are also linked to poor birth outcomes, impaired child cognitive development, poor nutritional status, and diminished early childhood well-being (14-18). Addressing these disorders is therefore essential to safeguarding both maternal and child health. A key step in this process is identifying and understanding their key predictors or risk factors, which is critical for designing targeted interventions.

Several studies have identified factors influencing depression in pregnant women, including low education, young maternal age, smoking during pregnancy, poor economic status, marital issues, and lack of social support (17, 19-21). However, findings remain inconsistent. For example, Muraca et al. found that younger maternal age was associated with maternal depression, while Agnafors et al. identified advanced maternal age as a significant risk factor (22, 23). Moreover, most of these studies have relied on traditional statistical models, which struggle to capture nonlinearity and interactions between variables, potentially leading to misinterpretations or inaccurate conclusions in complex datasets. Additionally, most studies that have employed advanced methods are from developed countries, highlighting a gap in the literature regarding low-resource settings.

Artificial intelligence (AI) techniques, specifically machine learning (ML) models, have shown promise in predicting mental health disorders such as depression, anxiety and post-traumatic stress disorder (24-28). These methods identify key predictors through importance rankings, inherently capturing nonlinear effects and complex interactions that conventional techniques often fail to address, thereby highlighting the most impactful points for intervention.

This study aimed to identify key predictors of maternal depression and anxiety in a low-resource setting using ML methods. The findings can inform the development and testing of targeted interventions to improve maternal mental health and promote healthy child development in similar contexts.

## Methods

This study was conducted to identify key predictors of MMH disorders, specifically anxiety and depression, among mothers of children aged 0 to 6 months in Kilifi, Kenya. The findings are intended to inform the development and testing of targeted interventions aimed at improving maternal well-being and child development. The following sections outline the specific procedures and techniques used throughout the research.

### Study setting

This study was conducted in Kaloleni and Rabai, two rural sub-counties in Kilifi County on the coast of Kenya, identified as some of the poorest regions in the country (29). The sub-counties cover an area of approximately 909 km² and have an estimated population of around 352,175 people, living in approximately 47,000 households. Both sub-counties have limited access to healthcare, with only 40 health facilities serving a large and scattered population. The area also experiences high levels of poverty, with approximately 70% of the population living below the national poverty line and 81% relying on subsistence agriculture, crafts, casual labour and petty trading for their livelihoods. In addition, the maternal health indicators for this area are well below national averages (30).

### Study participants

The study participants were 1,995 mothers of singleton children aged 0 to 6 months identified by Community Health Extension Workers (CHEWs) through the Kaloleni-Rabai Health and Demographic Surveillance System (KRHDSS), managed by the Department of Population Health (DPH) at Aga Khan University, Nairobi, Kenya (31). The KRHDSS is a population-based registry that covers all 47,000 households in the region and has been collecting data once a year since 2020 (31).

### Data collection

This study utilized baseline data collected between 10^th^ March 2023 and 28^th^ March 2024 as part of a broader longitudinal project titled “*Development and Validation of Artificial Intelligence (AI)/Machine Learning (ML)-Based Prediction Models for Poor Early Childhood Development in East Africa*.” The study enrolled 1,995 mothers of singletons children aged 0 to 6 months. Data collection occurred in participants’ households, identified by CHEWs in collaboration with Community Health Promoters (CHPs). Trained enumerators from Aga Khan University, aided by CHPs, conducted household visits to gather data. Mothers provided written informed consent for their own and their children’s participation in the study. Data were collected using tablets equipped with Open Data Kit, through interviewer-administered questionnaires as described below.

### Study tools

#### The 9-item Patient Health Questionnaire (PHQ-9)

The PHQ-9 (32) was utilized to assess maternal depressive symptoms. It includes 9 items rated on a Likert scale from 0 (not at all) to 3 (nearly every day). The total score, which ranges from 0 to 27, is calculated by summing individual item scores, with higher scores indicating greater levels of depressive symptoms. The PHQ-9 has been validated as a screening tool in studies conducted in Sub-Saharan African (SSA) countries (32-34).

#### The 7-item Generalised Anxiety Disorder (GAD-7)

The GAD-7 questionnaire (35) was used to assess maternal anxiety symptoms, including feelings of nervousness, worry, or restlessness over the past two weeks. It consists of 7 items rated on a Likert scale from 0 (not at all) to 3 (nearly every day). The total score, ranging from 0 to 21, is calculated by summing the individual item scores, with higher scores indicating greater levels of anxiety symptoms. The GAD-7 has been validated as a screening tool for generalized anxiety disorder (GAD) (35, 36).

#### Household Food Insecurity Access Scale (HFIAS)

The HFIAS questionnaire (37, 38) was used to assess household food insecurity levels. It includes 9 items evaluating aspects such as food supply uncertainty, insufficiency in the quality and quantity of food, going to bed hungry, and enduring full days and nights without eating over the past four weeks. The total score, calculated by summing the individual item scores, ranges from 0 to 27, with higher scores indicating greater levels of food insecurity. The HFIAS has been validated through various studies in developing countries (39-42).

#### Social demographic and economic questionnaire

The sociodemographic data collected for this study included age, marital status, religion, and other additional characteristics. Socioeconomic status was assessed using a 10-asset items commonly applied in the Kenyan context, which covers ownership of assets such as a radio, television, video machine, fridge/freezer, cooker, computer/tablet, bicycle, motorcycle, car/truck, and telephone (43). Each owned asset is assigned a value of 1, while the absence of an asset is scored as 0. In addition to assessing socioeconomic status, housing quality was evaluated based on specific characteristics, including the primary materials used for constructing floors (e.g., tiles, mud), walls (e.g., mud, bricks), and roofs (e.g., iron sheets, grass). Other factors considered included the type of toilet facilities (e.g., flush, bush) and the cooking area location (e.g., kitchen, outside the house), among others. These features were subsequently categorized into two classes: “improved” assigned a value of 1 or “unimproved” assigned a value of 0 (44, 45). A single wealth index score was calculated using the Multiple Correspondence Analysis (MCA) technique, which is particularly suited for binary data (46).

#### Health history questionnaire

A questionnaire with yes/no items was used to collect health history information for the mothers. The questionnaire addressed pregnancy-related complications, delivery challenges, and other relevant health factors.

#### Anthropometric measurements

Height and weight measurements were taken to calculate Body Mass Index (BMI), which was used as an indicator of maternal nutritional status. Measurements followed WHO-recommended procedures (47), and for quality control, each was taken three times.

### Ethics approval

The primary project was approved by the Aga Khan University, Nairobi Institutional Scientific and Ethics Review Committee (Ref: 2022/ISERC_75(V2)). Permission to conduct the study in Kenya was granted by the National Commission for Science, Technology, and Innovation (Ref: 2322698). A local permit to conduct the study was granted by the research office in Kilifi (Ref: KLF/DOH/RESEARCH/VOL.1/004). All participants provided written informed consent for their participation.

### Data analysis

#### Outcome variable

The primary outcomes of this study are maternal depression and anxiety symptoms, assessed using the PHQ-9 and GAD-7 scales, respectively. The PHQ-9 scores range from 0 to 27, with scores of 5–9, 10–14, and 15–27 indicating mild, moderate, and severe depressive symptoms (32). Similarly, GAD-7 scores range from 0 to 21, with scores of 5–9, 10–14, and 15–21 representing mild, moderate, and severe anxiety symptoms (35). Both outcomes were dichotomized into binary variables: scores ≥5 indicate the presence of symptoms (mild, moderate, and severe), while scores <5 indicate their absence (48, 49). The internal consistency of the scales in this study was acceptable, with Cronbach’s alpha values of 0.84 (95% CI: 0.83– 0.85) for PHQ-9 and 0.85 (95% CI: 0.84–0.86) for GAD-7.

#### Data preparation

The dataset used in this study consisted of 37 potential predictive features selected based on previous literature and domain expertise (S1 Table 1). Given the small number of missing values, imputation was performed using a combination of logical reasoning and statistical methods, including the mean, median, and mode, to ensure dataset consistency and accuracy. Data was processed and analysed using Python software (version 3.1.1) (50).

**Table 1.** Participant characteristics representing key predictors of depressive symptoms identified by ML models.

| Variable | Overall<br>N=1995 | Maternal depressive symptoms |  | P value |
| --- | --- | --- | --- | --- |
|  |  | No n (%) | Yes n (%) |  |
|  |  | 1693 (84.86%) | 302 (15.14%) |  |
| **Sociodemographic** |  |  |  |  |
| Age in years, median [Q1, Q3] | 27.0 [23.0, 32.0] | 27.0 [23.0, 32.0] | 28.0 [24.0, 33.0] | 0.006<sup>b</sup> |
| Number of pregnancies, median [Q1, Q3] | 3.0 [2.0,5.0] | 3.0 [1.0,5.0] | 3.0 [2.0,5.0] | <0.001<sup>b</sup> |
| Number of children, median [Q1, Q3] | 3.0 [1.0,4.0] | 3.0 [1.0,4.0] | 3.0 [2.0,5.0] | <0.001<sup>b</sup> |
| Number of children between 1 to 5 years, median [Q1, Q3] | 1.0 [0.0,1.0] | 1.0 [0.0,1.0] | 1.0 [0.0,1.0] | 0.179<sup>b</sup> |
| Marital status, n (%) |  |  |  | 0.001<sup>a</sup> |
| Married | 1770 (88.7) | 1520 (89.8) | 250 (82.8) |  |
| Unmarried | 225 (11.3) | 173 (10.2) | 52 (17.2) |  |
| Religion, n (%) |  |  |  | 0.095<sup>a</sup> |
| Christian | 1435 (71.9) | 1217 (71.9) | 218 (72.2) |  |
| Islam | 461 (23.1) | 400 (23.6) | 61 (20.2) |  |
| Traditional | 91 (4.6) | 70 (4.1) | 2 (0.7) |  |
| Other | 8 (0.4) | 6 (0.4) | 2 (0.7) |  |
| Caring for children with disability, n (%) |  |  |  | 0.018<sup>c</sup> |
| Yes | 31 (1.6) | 21 (1.2) | 10 (3.3) |  |
| No | 1964 (98.4) | 1672 (98.8) | 292 (96.7) |  |
| **Socioeconomic** |  |  |  |  |
| Wealth index, median [Q1, Q3] | -0.2 [-0.3,0.1] | -0.2 [-0.3,0.1] | -0.2 [-0.3,0.1] | 0.874<sup>b</sup> |
| **Health history** |  |  |  |  |
| Pregnancy complications, n (%) |  |  |  | <0.001<sup>a</sup> |
| Yes | 218 (10.9) | 157 (9.3) | 61 (20.2) |  |
| No | 1777 (89.1) | 1536 (90.7) | 241 (79.8) |  |
| Child survival in all pregnancies, n (%) |  |  |  | 0.018<sup>a</sup> |
| Yes | 1663 (83.4) | 1424 (84.1) | 239 (79.1) |  |
| No | 332 (16.6) | 269 (15.9) | 63 (20.9) |  |
| Household food insecurity scores, median [Q1, Q3] | 7.0 [1.0,13.0] | 6.0 [0.0,11.0] | 13.0 [8.0,17.0] | <0.001<sup>b</sup> |
| **Nutrition** |  |  |  |  |
| Maternal BMI, median [Q1, Q3] | 21.2 [19.3,24.0] | 21.2 [19.4,24.0] | 20.9 [18.7,23.7] | 0.056<sup>b</sup> |

a refers to Pearson's $\chi^2$ , b refers to the Kruskal-Wallis test, and c refers to Fisher's exact tests

#### Exploratory data analysis

The maternal characteristics were summarized using descriptive statistics, including proportions (%) and medians with their corresponding first and third quartiles. Group differences in participant characteristics were compared using Pearson’s χ² and Fisher’s exact tests for categorical data, and the Kruskal-Wallis test for data with a non-normal distribution. Hypothesis tests were conducted with a two-tailed approach, and statistical significance was set at p-value < 0.05. For continuous features, Pearson correlation coefficients were computed and plotted in a pairwise manner for all features identified as important or tentative. Decisions on feature retention were based on expert input, and the effect on predictive performance. Similarly, for categorical features, Cramér’s V correlation coefficients were calculated and plotted pairwise, and feature retention was guided by expert knowledge. Categorical features were encoded using the one-hot encoding method to allow easier interpretability of model outputs, S1 Table 2.

**Table 2.** Participant characteristics representing key predictors of anxiety symptoms identified by ML models.

| Variable | Overall | Maternal anxiety symptoms |  | P value |
| --- | --- | --- | --- | --- |
|  |  | No n (%) | Yes n (%) |  |
|  |  | 1822 (91.33%) | 173 (8.67%) |  |
| **Sociodemographic** |  |  |  |  |
| Age in years, median [Q1, Q3] | 27.0 [23.0, 32.0] | 27.0 [23.0, 32.0] | 28.0[24.0,34.0] | 0.004<sup>b</sup> |
| Number of pregnant, median [Q1, Q3] | 3.0 [2.0, 5.0] | 3.0 [1.0, 5.0] | 3.0 [2.0, 5.0] | 0.007<sup>b</sup> |
| Number of children, median [Q1, Q3] | 3.0 [1.0, 4.0] | 3.0 [1.0, 4.0] | 3.0 [2.0, 5.0] | 0.008<sup>b</sup> |
| Number of children between 1 and 5 years, median [Q1, Q3] | 1.0 [0.0, 1.0] | 1.0 [0.0, 1.0] | 1.0 [0.0, 1.0] | 0.365<sup>b</sup> |
| Religion, n (%) |  |  |  | 0.391<sup>a</sup> |
| Christian | 1435 (71.9) | 1302 (71.5) | 133 (76.9) |  |
| Islam | 461 (23.1) | 430 (23.6) | 31 (17.9) |  |
| Traditional | 91 (4.6) | 83 (4.6) | 8 (4.6) |  |
| Other | 8 (0.4) | 7 (0.4) | 1 (0.6) |  |
| Caring for children with disability, n (%) |  |  |  | 0.183<sup>c</sup> |
| Yes | 31 (1.6) | 26 (1.4) | 5 (2.9) |  |
| No | 1964 (98.4) | 1796 (98.6) | 168 (97.1) |  |
| **Socioeconomic** |  |  |  |  |
| Wealth index, median [Q1, Q3] | -0.2 [-0.3,0.1] | -0.2 [-0.3,0.1] | -0.1 [-0.3,0.2] | 0.598<sup>b</sup> |
| Education status, n (%) |  |  |  | 0.826<sup>a</sup> |
| None (No formal education) | 317 (15.9) | 288 (15.8) | 29 (16.8) |  |
| Formal education | 1678 (84.1) | 1534 (84.2) | 144 (83.2) |  |
| **Health history** |  |  |  |  |
| Pregnancy complication, n (%) |  |  |  | <0.001<sup>a</sup> |
| Yes | 218 (10.9) | 182 (10.0) | 36 (20.8) |  |
| No | 1777 (89.1) | 1640 (90.0) | 137 (79.2) |  |
| Place of delivery, n (%) |  |  |  | 0.508<sup>a</sup> |
| Home | 218 (10.9) | 196 (10.8) | 22 (12.7) |  |
| Hospital/clinic | 1777 (89.1) | 1626 (89.2) | 151 (87.3) |  |
| Child survival in all pregnancies, n (%) |  |  |  | 0.012<sup>a</sup> |
| Yes | 1663 (83.4) | 1531 (84.0) | 132 (76.3) |  |
| No | 332 (16.6) | 291 (16.0) | 41 (23.7) |  |
| Attended antenatal clinic, n (%) |  |  |  | 0.017<sup>a</sup> |
| Yes | 1901 (95.3) | 1743 (95.7) | 158 (91.3) |  |
| No | 94 (4.7) | 79 (4.3) | 15 (8.7) |  |
| **Household food insecurity scores, median [Q1, Q3]** | 7.0 [1.0, 13.0] | 6.0 [1.0, 12.0] | 13.0 [8.0, 17.0] | <0.001<sup>b</sup> |
| **Nutrition** |  |  |  |  |
| Maternal BMI, median [Q1, Q3] | 21.2 [19.3, 24.0] | 21.2 [19.4, 24.0] | 21.1 [18.9,24.1] | 0.382<sup>b</sup> |

a refers to Pearson's $\chi^2$ , b refers to the Kruskal-Wallis test, and c refers to Fisher's exact tests

#### Model development

This study investigated three supervised ML models (classifiers): Random Forest (RF), Logistic Regression (LR) with L2 regularisation (Ridge), and Extreme Gradient Boosting (XGBoost). These algorithms were chosen for their efficiency, widespread application in healthcare research, and their potential for explainability (51, 52).

The model development process began with stratifying the dataset by the outcome variable to ensure balanced representation in both the training and test sets. Following the commonly used 80:20 data splitting ratio (53), the data was randomly divided into 80% training and 20% testing for both outcomes (anxiety and depressive symptoms).

To address the issue of class imbalance efficiently, class weights were adjusted using the *class_weight=’balanced’* parameter in the RF and LR algorithms (54). This method automatically assigns weights inversely proportional to class frequencies in the input data, promoting fairness in the model’s predictions. In contrast, for the XGBoost model, the *scale_pos_weight* parameter was utilized, calculated as the ratio of the positive class proportion to the negative class proportion (55). This adjustment ensures that the model gives appropriate emphasis to the minority class, enhancing its performance in imbalanced datasets.

To fine-tune the models and select the best hyperparameters, 10-fold cross-validation was applied to the training set. This involved dividing the training data into 10 equal parts. At each fold, 90% of the training data was used for model training, while the remaining 10% was used for validation (56).

Grid search was used for hyperparameter tuning to identify the optimal model, with performance evaluated through 10-fold cross-validation as mentioned above and the area under the receiver operating characteristic curve (AUC). This metric provided insights into each classifier’s ability to distinguish between positive and negative classes, particularly in scenarios of class imbalance (57).

#### Feature selection

The feature selection process was based on feature importance rankings derived from SHapley Additive exPlanations (SHAP) values, as proposed by Lundberg et al. (58). The SHAP method offers a more consistent method for calculating feature importance and effects compared to traditional methods like gain-based importance in ensemble tree models. Based on game theory, SHAP fairly distributes the value among features based on their contribution to the model’s predictions.

#### Model performance evaluation

After obtaining the optimal model, its performance was evaluated using the remaining 20% of the dataset for both outcomes, which had been reserved for testing. These test sets provided an evaluation of how well the model would generalize to unseen data, as they had not been utilized in the training or validation phases. Various performance metrics, including accuracy, sensitivity, and specificity, were employed to gain a comprehensive understanding of the model’s performance (59). The Youden Index was employed to identify the optimal classification cutoff (60). Additionally, the AUC was calculated to assess the model’s ability to discriminate between different outcome classes across various decision thresholds (61). This final evaluation on the test sets constitutes an internal validation, given that the test data is drawn from the same original dataset.

#### Model interpretation

The explainability of the best-performing models on internal validation was explored by examining feature importance and feature effects using SHAP values. Two SHAP methods were used: SHAP dependence plots, which visualize the relationship between individual features and the model’s predictions, and SHAP summary plots (bar plots and beeswarm plots), which display feature importance. Both types of plots were generated using the test data to assess feature importance and effect.

## Results

Tables 1 and 2 present key maternal characteristics of the 1,995 mothers included in the modelling process, highlighting predictors for depressive (Table 1) and anxiety (Table 2) symptoms. The median age of participants was 27 years (Q1: 23.0, Q3: 32.0). Among the mothers, 302 (15.14%) exhibited depressive symptoms (mild, moderate, and severe), while 173 (8.67%) experienced anxiety symptoms (mild, moderate, and severe).

Significant group differences were observed in most of the characteristics for both outcomes, including sociodemographic, socioeconomic, health history, food insecurity, and nutrition factors (p-value < 0.05; Tables 1 and 2). S1 Figs 1 and 2 present correlation matrices for both numerical and categorical features. Among the numerical features, the number of children and the number of pregnancies were highly correlated (r=0.96). Both were retained in the modeling process based on domain expert advice, as they capture distinct aspects of reproductive history: the number of children represent the total number of living children, while the number of pregnancies encompasses the full pregnancy history, including live births and miscarriages. Similarly, although living with a partner and marital status showed a strong correlation (r = 0.87), and place of delivery and delivery assistant were also highly correlated (r = 0.97), both pairs were retained based on expert guidance.

**Fig 1.**
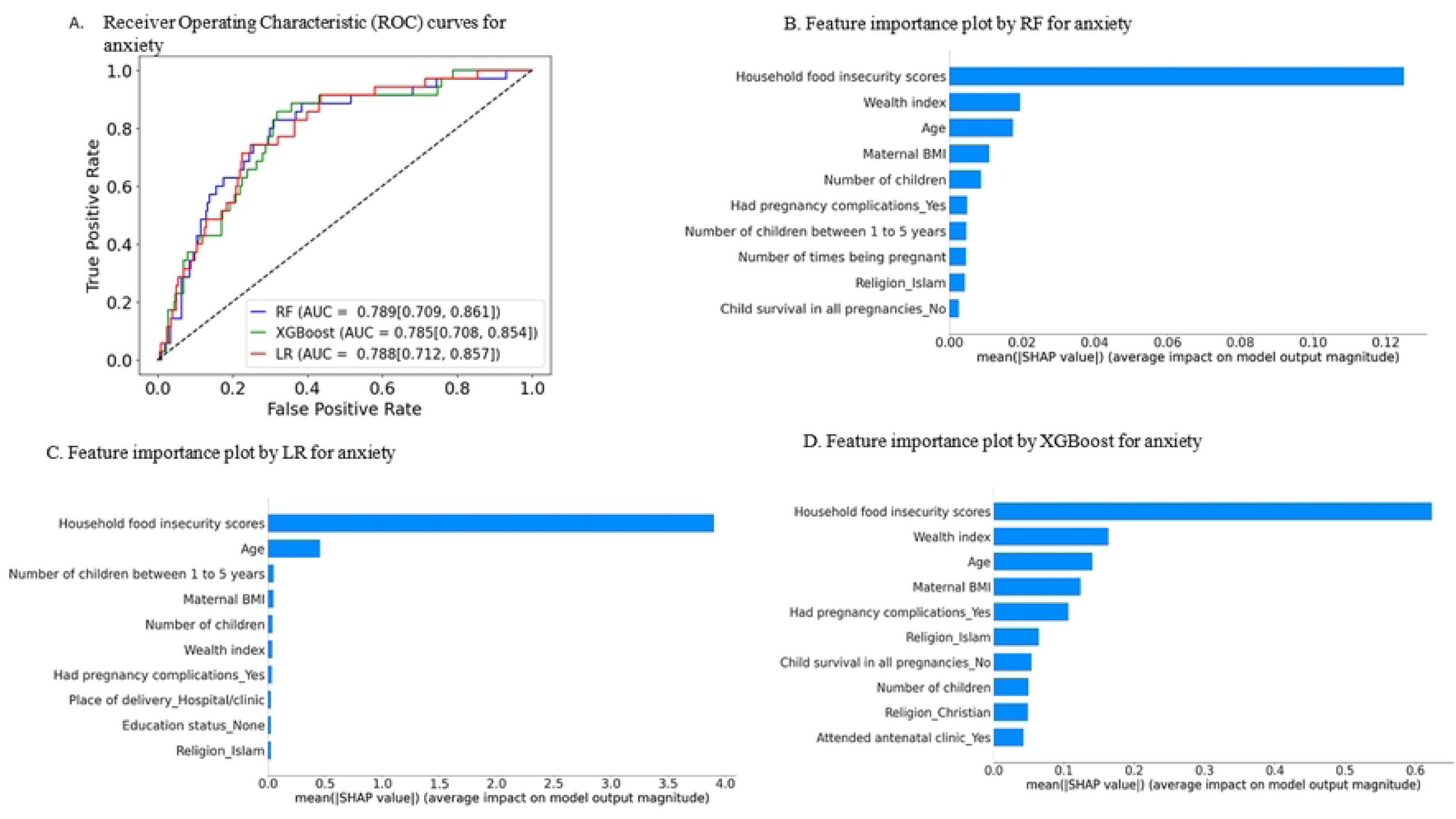
Plot A presents the Receiver Operating Characteristic (ROC) curves for three models, Random Forest (RF), Extreme Gradient Boosting (XGBoost), and Logistic Regression (LR) along with their corresponding AUC values for predicting anxiety symptoms. SHAP feature importance plots (B–D) display the top 10 important predictors influencing anxiety symptoms, ranked from most to least important.

**Fig 2.**
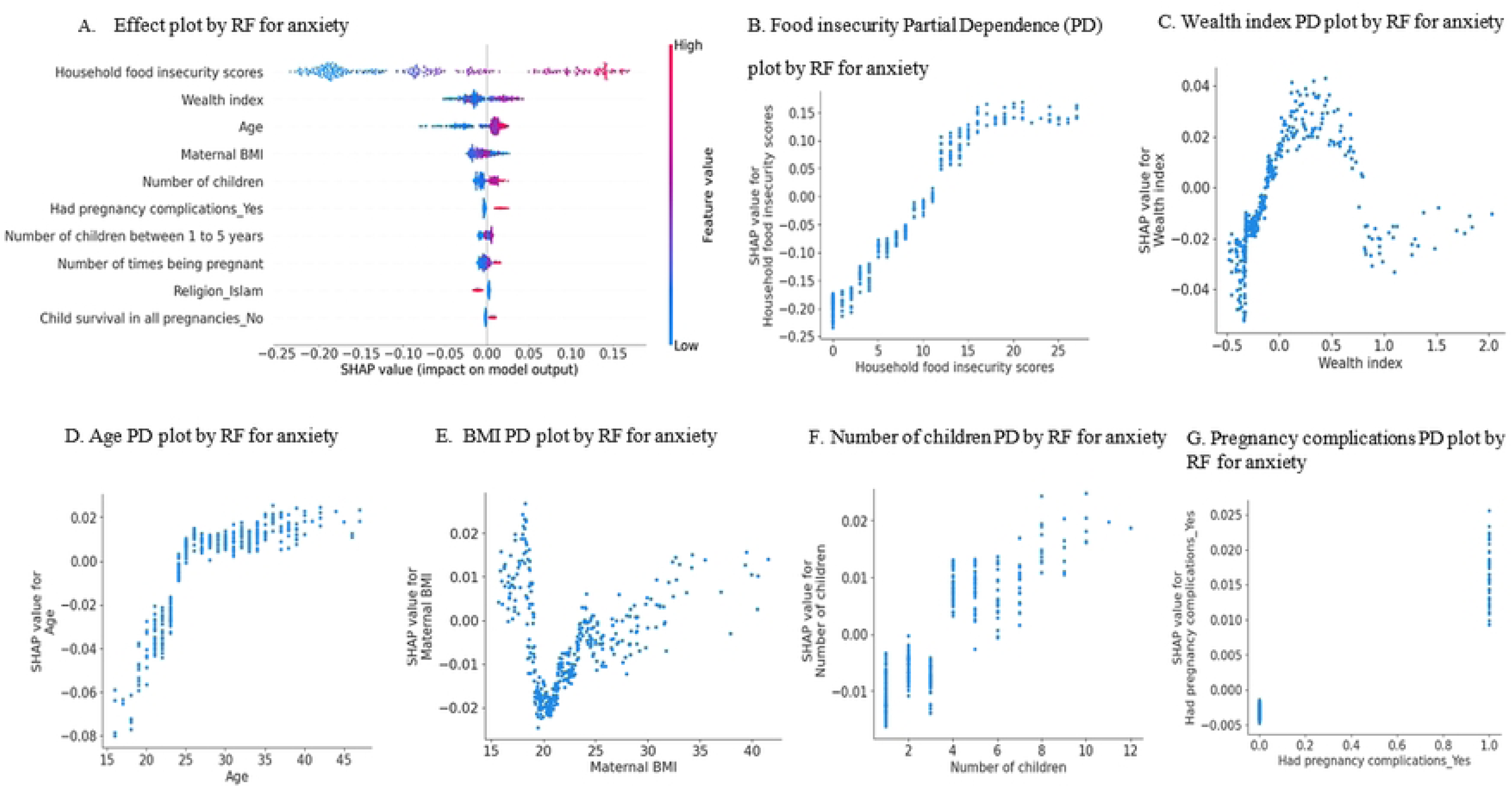
Plot A, an effect plot, (SHAP beeswarm plot) shows the top 10 important predictors influencing anxiety symptoms from a Random Forest (RF) model, ranked by importance. Color indicates the feature value (red = high, blue = low), and position along the x-axis shows the direction and strength of impact. Plots B–G present SHAP partial dependence for the top 6 key predictors: increased food insecurity, age, and number of children, as well as low wealth index, low BMI, and pregnancy complications, all of which have a positive impact on anxiety symptoms.

### Model performance evaluation

Table 3 summarizes the performance of all three models, each achieving AUCs above 0.70 on unseen test data using 23 features. In predicting anxiety symptoms, the RF model achieved the highest AUC at 0.789 (95% CI: 0.709–0.861), followed closely by logistic regression at 0.788 (95% CI: 0.712–0.857) and XGBoost at 0.785 (95% CI: 0.708–0.854), with the differences being statistically insignificant. Additional performance metrics, including accuracy, sensitivity, and specificity, are also presented in the table. When using a reduced set of the 10 most important features selected via the SHAP method, model performance remained stable. RF achieved an AUC of 0.784 (95% CI: 0.701–0.855), with LR at 0.786 (95% CI: 0.708–0.859) and XGBoost at 0.792 (95% CI: 0.709–0.867).

**Table 3.** Evaluation of model performance on unseen test data.

| Outcome variable | Algorithm | No of features | Test size | Best cutoff | Sensitivity (95%CI) | Specificity (95%CI) | AUC (95%CI) | Accuracy (95%CI) |
| --- | --- | --- | --- | --- | --- | --- | --- | --- |
| Anxiety | Logistic regression | 23 | 399 (20%) | 0.497 | 0.743 [0.738 - 0.748] | 0.701 [0.699 - 0.702] | **0.788 [0.712, 0.857]** | 0.704 [0.703 - 0.706] |
|  |  | 10 | 399 (20%) | 0.490 | 0.714 [0.710 - 0.719] | 0.684 [0.683 - 0.686] | 0.786 [0.708, 0.859] | 0.687 [0.685 - 0.688] |
|  | XGBoost | 23 | 399 (20%) | 0.520 | 0.657 [0.652 - 0.662] | 0.739 [0.738 - 0.741] | **0.785 [0.708, 0.854]** | 0.732 [0.730 - 0.733] |
|  |  | 10 | 399 (20%) | 0.529 | 0.714 [0.710 - 0.719] | 0.736 [0.735 - 0.738] | 0.792 [0.709, 0.867] | 0.734 [0.733 - 0.736] |
|  | Random forest | 23 | 399 (20%) | 0.491 | 0.743 [0.738 - 0.748] | 0.728 [0.727 - 0.730] | **0.789 [0.709, 0.861]** | 0.729 [0.728 - 0.731] |
|  |  | 10 | 399 (20%) | 0.466 | 0.686 [0.681 - 0.691] | 0.739 [0.738 - 0.741] | 0.784 [0.701, 0.855] | 0.734 [0.733 - 0.736] |
| Depression | Logistic regression | 23 | 399 (20%) | 0.486 | 0.633 [0.629 - 0.637] | 0.711 [0.709 - 0.713] | **0.724 [0.656, 0.785]** | 0.699 [0.698 - 0.701] |
|  |  | 10 | 399 (20%) | 0.480 | 0.617 [0.613 - 0.621] | 0.693 [0.692 - 0.695] | 0.723 [0.652 - 0.783] | 0.682 [0.680 - 0.683] |
|  | XGBoost | 23 | 399 (20%) | 0.536 | 0.533 [0.529 - 0.537] | 0.717 [0.715 - 0.718] | **0.705 [0.628, 0.772]** | 0.689 [0.688 - 0.691] |
|  |  | 10 | 399 (20%) | 0.538 | 0.533 [0.529 - 0.537] | 0.717 [0.715 - 0.718] | 0.705 [0.630, 0.772] | 0.689 [0.688 - 0.691] |
|  | Random forest | 23 | 399 (20%) | 0.558 | 0.583 [0.579 - 0.587] | 0.714 [0.712 - 0.715] | **0.711 [0.642, 0.774]** | 0.694 [0.693 - 0.696] |
|  |  | 10 | 399 (20%) | 0.551 | 0.583 [0.579 - 0.587] | 0.723 [0.721 - 0.724] | 0.708 [0.638, 0.772] | 0.702 [0.700 - 0.703] |

In predicting depressive symptoms, LR, XGBoost, and RF demonstrated similar performance, with AUCs of 0.72 (95% CI: 0.656–0.785), 0.705 (95% CI: 0.628–0.772), and 0.711 (95% CI: 0.642–0.774) respectively. Detailed performance metrics such as accuracy, sensitivity, and specificity are also included in the table. When limited to the top 10 SHAP-selected features, the models maintained comparable performance, with AUCs of 0.723 (95% CI: 0.652–0.783) for LR, 0.705 (95% CI: 0.630–0.772) for XGBoost, and 0.708 (95% CI: 0.638–0.772) for RF.

### Model interpretation

Figs 1 and 3 present the Receiver Operating Characteristic (ROC) curves and feature importance plots generated using all 23 features. For both anxiety and depressive symptoms, household food insecurity scores consistently emerged as a critical predictor across all models. Other key predictors among the top 10 important features included sociodemographic factors such as number of children, age, number of pregnancies, marital status, religion, caring for children with disabilities, and number of children aged 1 to 5 years. Socioeconomic status indicators such as education status and wealth index, nutritional status measured by maternal BMI, and health history factors including pregnancy complications, place of delivery, antenatal care attendance, and child survival in all pregnancies. Figs 2 and 4 illustrate the magnitude, direction of effects, and impact of these features on the prediction of depressive and anxiety symptoms.

**Fig 3.**
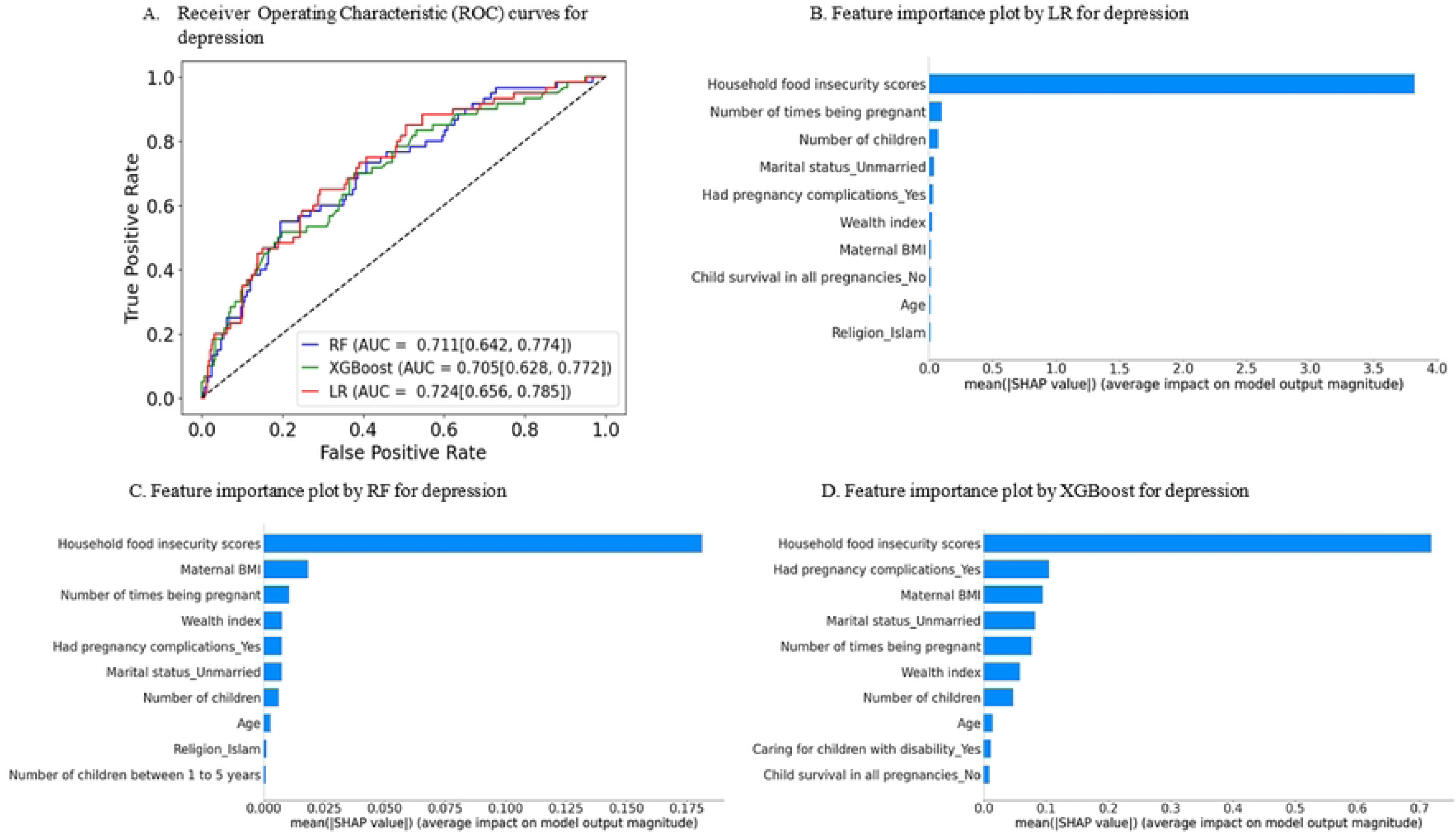
Plot A presents the Receiver Operating Characteristic (ROC) curves for three models, Random Forest (RF), Extreme Gradient Boosting (XGBoost), and Logistic Regression (LR) along with their corresponding AUC values for predicting depressive symptoms. SHAP feature importance plots (B–D) display the top 10 important predictors influencing depressive symptoms, ranked from most to least important.

**Fig 4.**
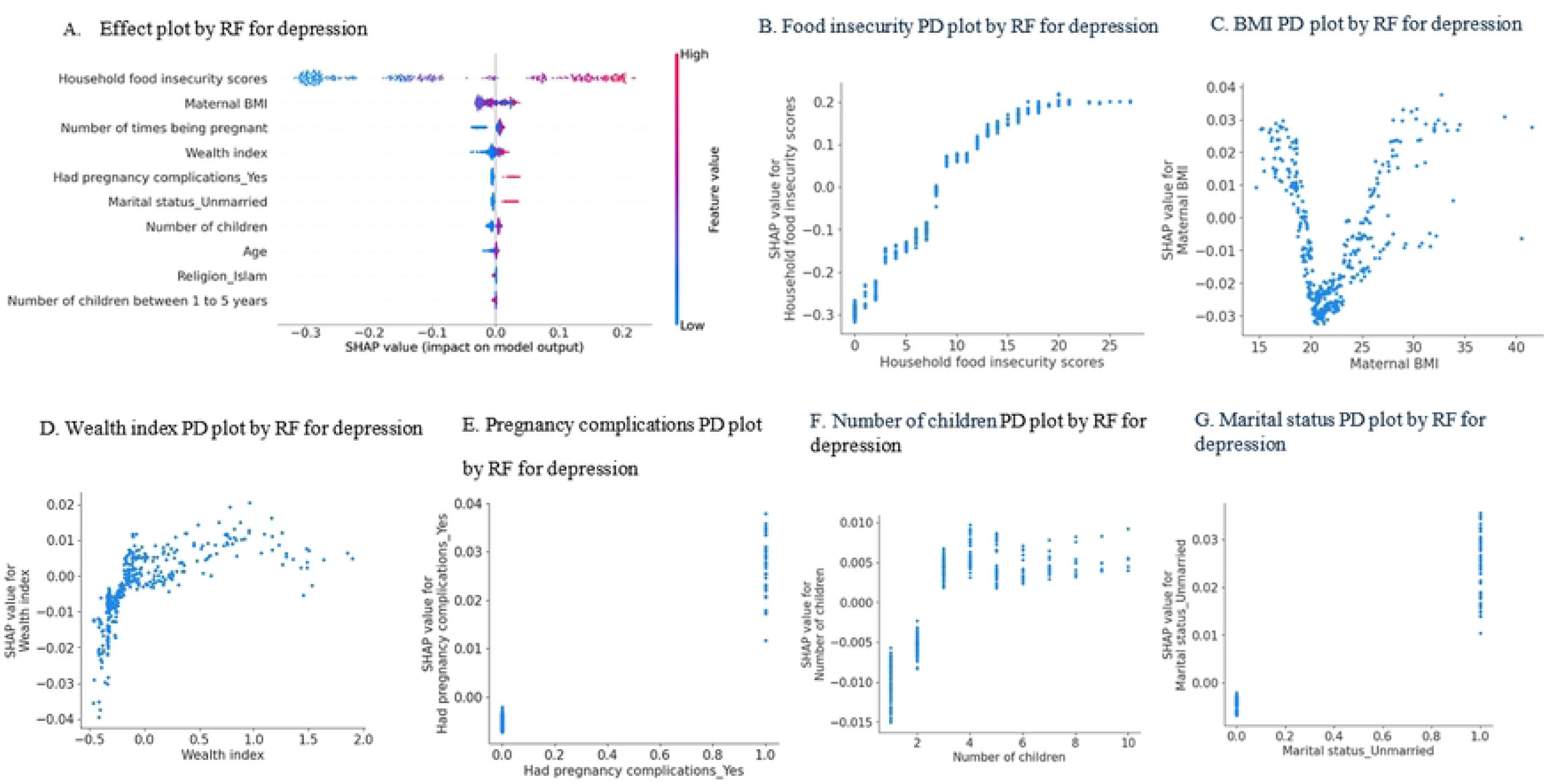
Plot A (SHAP beeswarm plot) shows the top 10 important predictors influencing depressive symptoms from a Random Forest (RF) model, ranked by importance. Color indicates the feature value (red = high, blue = low), and position along the x-axis shows the direction and strength of impact. Plots B–G present SHAP partial dependence for the top 6 key predictors: increased food insecurity, number of children, as well as low BMI, low wealth index, pregnancy complications and unmarried marital status, all of which have a positive impact on depressive symptoms.

## Discussion

This study investigated the key predictors of MMH disorders, specifically anxiety and depression, among mothers of children aged 0 to 6 months, using ML techniques. The findings highlight the mental health burden in low-resource settings and underscores critical areas for targeted interventions.

The study found that 15.14% of mothers exhibited mild to severe symptoms of depression, and 8.67% showed mild to severe symptoms of anxiety. These results are consistent with findings from LMICs, where maternal anxiety and depression are prevalent, with reported rates ranging from 10% to 25% for depression and 8% to 16% for anxiety during pregnancy and the postpartum period (6, 8). However, studies in SSA have reported even higher prevalence rates, with depression ranging from 10% to 70% and anxiety from 10% to 50% (9-13). The variation in prevalence across studies may be due to differences in screening tools, sample characteristics, and other methodological or population related factors. The observed prevalence underscores the need for mental health services for pregnant and postpartum women in low-resource settings.

Three ML models (LR, XGBoost and RF) were employed to predict maternal depression and anxiety using both the full set of 23 features and a reduced subset of 10 features selected through SHAP. For both depression and anxiety, all models achieved AUC values above 0.70, indicating good discriminative performance in identifying mothers with and without symptoms (62). Although the differences in AUC were not statistically significant, RF achieved the highest AUC of 0.789 for anxiety prediction, while LR performed slightly better for depression prediction with an AUC of 0.724. These findings are consistent with previous studies where machine learning classifiers reached AUCs above 0.70 for predicting depression in pregnant and postpartum women (63-65). All models also showed comparable performance across other metrics, including accuracy, sensitivity, and specificity. Interestingly, the models predicted anxiety more accurately than depression using the same feature set, which may be due to the selected features being more predictive of anxiety. Notably, model performance remained stable when using only the top 10 SHAP-selected features, suggesting that accurate predictions can be achieved even with a reduced feature set. This study also suggests that a simple LR model can achieve good results with a comparable sample size. However, more complex models, such as RF and XGBoost, may be more effective when feature interactions are nonlinear, as they can better capture these complex relationships.

To identify key predictors, the 10 most important features from each model were selected, revealing both overlapping and unique risk factors for maternal depression and anxiety. Consistently across all models, household food insecurity, wealth index, age, maternal BMI, number of children, and pregnancy complications emerged as overlapping predictors for both conditions. Additional unique predictors identified across models included the number of children aged 1 to 5 years, unmarried status, child survival in all pregnancies, antenatal clinic attendance, religion, lack of formal education, place of delivery (hospital or clinic), and caring for children with disabilities. Although religion emerged as one of the key predictors, its influence may not be entirely independent. Interactions between religion (specifically, Christian and Islam) and other key predictors such as food insecurity, wealth index, and maternal BMI were explored (S1 Fig 3). The interactions involving the Christian religion appeared stronger than those involving Islam, suggesting that religion may act as a modifying or confounding factor, potentially shaping how other key predictors relate to MMH outcomes. These findings align with existing literature, with studies using traditional statistical methods highlighting the significant role of sociodemographic, socioeconomic, nutritional, and health-related factors in contributing to poor mental health outcomes (22, 66-71). Notably, household food insecurity consistently emerged as a critical predictor for both depression and anxiety, underscoring its importance as a primary target for intervention strategies. Other overlapping predictors should also be considered as priorities in intervention planning following food insecurity.

This study employed robust methodological approaches, including validated screening tools (PHQ-9 and GAD-7) and advanced techniques (ML models) for predictive modeling. Our findings suggest that ML techniques can be effectively used to screen pregnant and postpartum women in low-resource settings, identifying those at highest risk of mental health issues and facilitating the development of targeted preventive interventions. The use of SHAP values for feature importance and effect analysis enhanced model interpretability, providing actionable insights for policymakers and practitioners.

However, this study has some limitations, including a small sample size and an imbalanced dataset, which may restrict the predictive power of the ML models. Additionally, data collection via questionnaires during interviews could have introduced bias, particularly underreporting of symptoms due to stigma. A larger, longitudinal study is needed to collect data at multiple time points for external validation and to further strengthen the findings, as the focus on mothers of children aged 0 to 6 months limits the generalizability to other maternal populations.

## Conclusion

In conclusion, this study provides valuable insights into the prevalence and key predictors of maternal depression and anxiety among mothers of children aged 0 to 6 months in a low-resource setting. The findings highlight the significant mental health burden faced by mothers in such environments, with factors such as household food insecurity, wealth index, age, maternal BMI, number of children, and pregnancy complications identified as key predictors for both conditions. The use of ML techniques demonstrated the potential for identification of high-risk individuals, enabling targeted interventions to address maternal mental health issues. Notably, food insecurity emerged as a critical predictor of both depression and anxiety, emphasizing the need for focused interventions in this area. These findings provide critical evidence for integrating food security measures into mental health interventions and policies targeting maternal populations. While the study’s small sample size limits the generalizability of the results, it paves the way for further research with larger, longitudinal datasets to validate these findings and refine predictive models for maternal mental health in low-resource settings.

## Data Availability

The data that support the findings of this study are available from the corresponding authors upon request. Due to ethical and confidentiality considerations, the data are not publicly available.

## Acknowledgement

We would like to thank all the participants who voluntarily took part in this study. We are also grateful to the Kilifi County health management team, the Kaloleni and Rabai Sub-counties health management teams, community health extension workers, and community health promoters from both sub-counties for their invaluable support throughout the study. We further acknowledge Anne Muthoki Mutua, Asha Mnyazi Tunje, Erick Kopo Mataza, Faith Pola Karisa, Bath Ochieng Ochieng, and Bilal Masika for their significant contributions to data collection. Special thanks to Candace Kolars and Eileen Haus for reviewing the manuscript.

## Supporting information

S1 Table 4. Features considered relevant for modelling process indicating few missing values.

S1 Fig 5. Correlation between numerical features.

S1 Fig 6. Correlation between categorical features

S1 Table 5. Features retained for modelling process after one hot encoding.

S1 Fig 7. Plots illustrating the interactions between religion and key predictors (food insecurity, wealth index, and maternal BMI).

